# The proportion testing positive for SARS-COV-2 among the tested population in the U.S.: Benefits of the positive test ratio under scaled testing scenarios

**DOI:** 10.1101/2020.04.21.20074070

**Authors:** George Ng, Constance Wang

**Affiliations:** University of California, Berkeley, School of Public Health; University of California, Berkeley, College of Engineering

**Author notes:** Corresponding author: George Ng, PhD, University of California, Berkeley, School of Public Health, 2121 Berkeley Way West Berkeley, CA 94720-1760.

## Abstract

The ratios offer simple ways to account for variations in testing and reporting. Tracking the ratios in addition to cases offer a more precise view of the pandemic. Our observations underscore the need to scale mass testing with accurate and reliable tests, to implement testing systematically and report results consistently.

---

What clinicians and public health experts need most in the next phase of COVID-19 response is an estimate of the prevalence of SARS-COV-2 in the US population.^1-2^ However, in this period before researchers obtain evidence from population-based studies on infection dynamics with accurate and reliable tests, it is still critical to have an idea of the proportion testing positive for infection among the tested population, even given variations in test accuracy, reliability and differences in testing criteria over time.

Tracking the number of confirmed positives each day over time for a city or state^3^ is problematic since this number (numerator) lacks a defined denominator for estimation. Fluctuations in the number of confirmed cases may reflect uneven testing capacity, batched processing or reporting. A city may appear to have more cases just from ramping up testing, or may appear to have a resurgence or reduction of cases due to changes in who can get tested, availability of test supplies, federal funding for testing sites and other shifts in regulator processes.

Estimating the proportion testing positive has been difficult. Significant time lags exist from samples collection to processing and again, to results reporting. Additionally, we lack an official, publicly-available centralized platform that compiles and links day-to-day data for each city or state on the number tested, processed, and number of positives.

## Methods

We evaluated the usefulness of the positive test ratio and the cumulative positive test ratio to account for the uneven number of tests processed or results reported across U.S. states from March 23-April 12. The positive ratio is the number of positives divided by the number of results reported for a given date; the cumulative ratio is the cumulative number of positives divided by the cumulative number of results reported through that date. We used data from the COVID-19 tracking project, a public platform that compiles and links data on test results from official sources in the US.^4^ The cumulative percentage of the population tested for each state by date was calculated using data from the 2019 U.S. Census.

## Results

Figure 1 presents the cumulative ratio by percentage of the population tested for 15 U.S. states with the highest number of confirmed cases. When the testing strategy moves beyond close contacts, the cumulative ratio stabilizes to give an indication of the prevalence of infection among the tested population.

**Figure 1.**
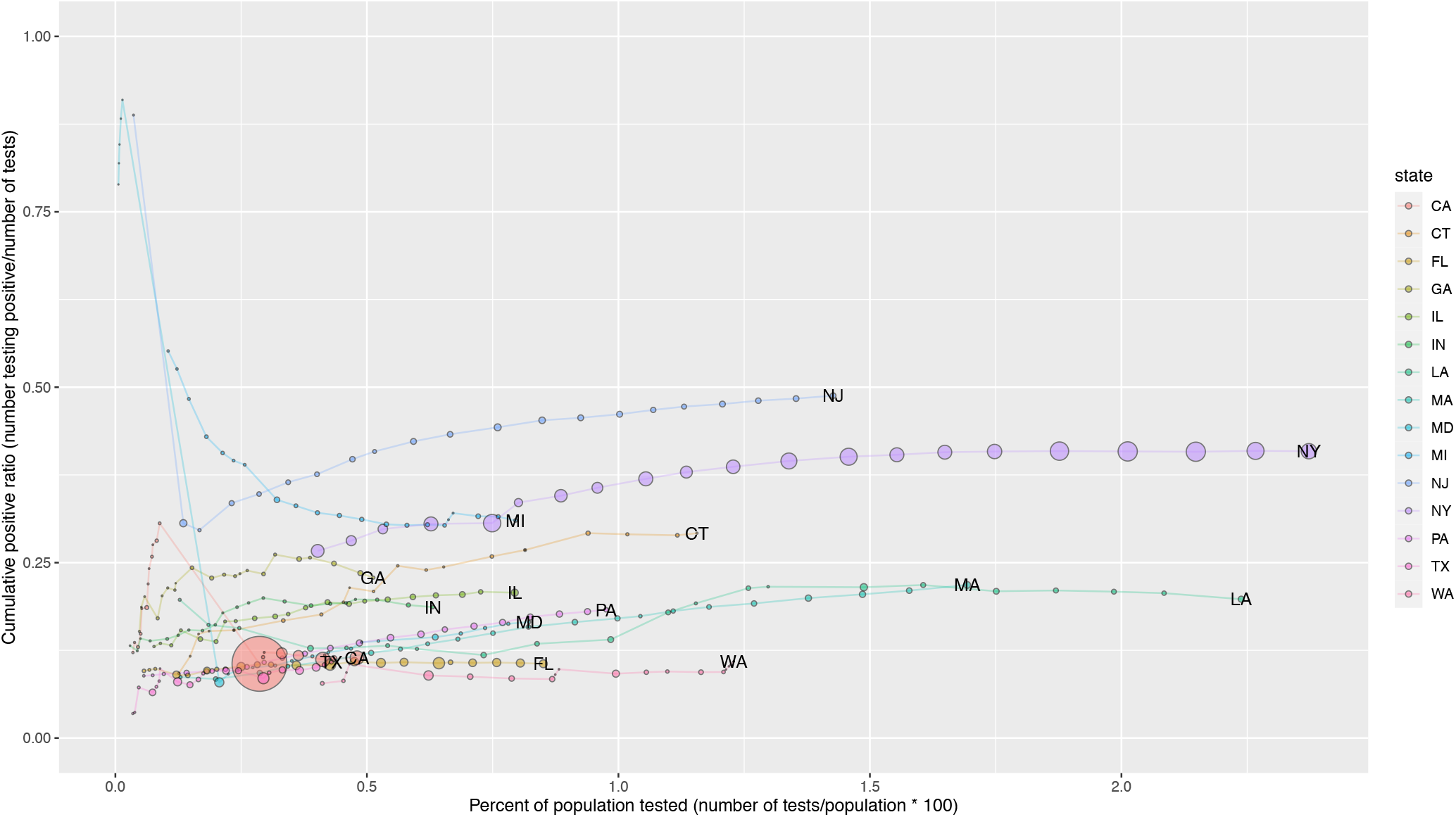
Cumulative positive test ratio and percentage of the population tested for 15 US states with the highest number of COVID-19 confirmed cases.

The cumulative ratios are higher initially, as expected, due to restricted testing of close contacts of confirmed cases. When a progressively higher proportion of the population is tested, the cumulative ratio converges, such as the ratios of New York and Washington state. In New York, New Jersey and Connecticut, once there was more testing, it became clear the prevalence was higher among the tested than the initial period. Among these 15 states, California ranks among the lowest in testing with the most inconsistencies in results reporting.

The day-to-day positive ratio is sensitive to systematic differences, such as in how the tests are batched and reported (Figure 2). If a given day’s reports are all from a senior facility with significant risk of transmission, then the ratio on that day could be higher than that of another day. On the other hand, if the day-to-day positive ratio is consistently decreasing over a period, this could be evidence of flattening the curve.

**Figure 2.**
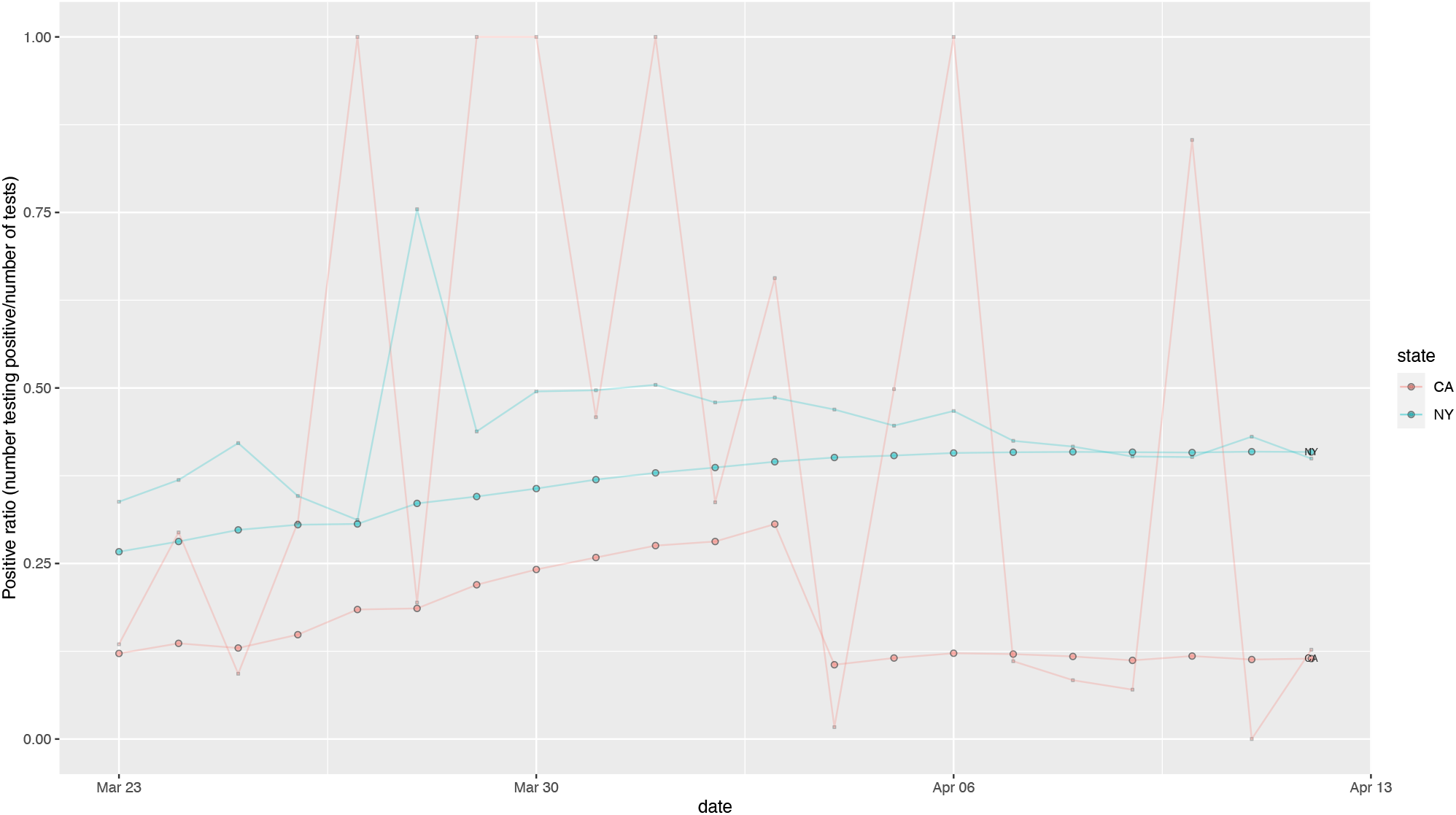
Day-to-day positive test ratio and cumulative positive test ratio by reporting day in New York and California.

## Data Availability

The COVID Tracking Project

https://covidtracking.com/

## Author Contributions

Drs. Ng and Wang had full access to all the data in the study and take responsibility for the integrity of the data and the accuracy of the data analysis. Drs. Ng and Wang contributed equally to the concept and design, analysis, interpretation and drafting of the manuscript.

## Conflict of Interest Disclosures

None reported.

## Notes

### Competing Interest Statement

The authors have declared no competing interest.

### Funding Statement

No payments received

